# Dynamics of serum BDNF After Aerobic Exercise in Subacute Stroke Patients: A Sub-Analysis of the randomized controlled Phys-Stroke trial

**DOI:** 10.1101/2024.09.24.24314322

**Authors:** Torsten Rackoll, Lea Doppelbauer, Konrad Neumann, Alexander Heinrich Nave, the Phys-Stroke study group

## Abstract

During the very early phase of stroke recovery, the brain is hypothesized to be in a neuroplastic state that is responsive to external stimuli. Here, brain-derived neurotrophic factor (BDNF) is thought to play an important role. BDNF, a member of the neurotrophin family, is implicated in processes such as synaptogenesis and long-term potentiation. Post-stroke, reduced BDNF levels are observed compared to the healthy population. An intervention that has been shown to elevate BDNF is Aerobic exercise. Yet, the impact of aerobic exercise in stroke patients remains unclear. This sub-analysis of the multicenter, randomized, controlled ’Physical Fitness Training in Patients with Subacute Stroke’ trial aimed to investigate serum BDNF dynamics and the effects of a four-week aerobic exercise intervention on long-term BDNF levels.

Data from 200 patients, with missing data imputed, revealed a modest increase in serum BDNF levels up to three months post-stroke (22.6 ng/ml, 95% CI 19.2 to 26) which plateaued until six months (24.3 ng/ml, 95% CI 20.6 to 27.9). Despite higher baseline BDNF levels in the exercise group, no treatment effect was observed until six months (training 24.6 ng/ml, 95% CI 20.2 to 29 vs relaxation: 24.3 ng/ml, 95% CI 19.8 to 28.8, *p* = 0.95). A sex-related interaction was identified in the relaxation group, with female patients exhibiting higher BDNF increases until end of follow up (20.9 ng/ml, 95% CI 15.7 to 26.2 (males) vs 31.0 ng/ml, 95% CI 22.9 to 39.0 (females), *p* = 0.039). Dose-response analyses and associations with recurrent events yielded no substantial differences.

While serum BDNF levels increased early after stroke in the first three months, aerobic exercise did not demonstrate a significant impact on BDNF levels within this dataset. Further investigations with more measurements during the early subacute phase are warranted to elucidate the interplay between exercise, BDNF, and recovery.

## Introduction

The early phase after stroke is hypothesized as a critical time window as the brain responds to the structural damage with increasedendogenous neuroplasticity. Most of the functional recovery is argued to occur in the early weeks after stroke.(1,2) Within this timeframe, processes of rewiring of structural and functional brain connections are activated and new neurons can evolve in certain brain areas.(3) Brain derived neurotrophic factor (BDNF) is part of the neurotrophin family and is involved in neuroplasticity.(4) It is primarily synthesized in the endoplasmic reticulum and is said to be able to cross the blood brain barrier. Although about 75% of BDNF are synthesized in the brain, BDNF is internalized and transported in platelets and therefore serum BDNF is regarded as a potential proxy of circulating BDNF in the brain.(5–7) Mature BDNF, the end product of BDNF synthesis, binds to the tyrosin-related kinase receptor B (TrkB) and via this pathway is involved in neuronal survival, synaptogenesis, outgrowth of dendritic spines and long-term potentiation.(8) In the brain, highest level of BDNF can be found in the CA3 region of the Hippocampus as well as the dentate gyrus, the amygdala and some cortical areas such as the visual and somatosensory cortex. Reduced levels of BDNF have been consistently shown in patients with cognitive impairments, highlighting its role in memory formation.(9)

In the stroke population, BDNF has been shown to be decreased compared to healthy individuals, and BDNF levels remained unchanged up to more than a month post stroke.(10) Further, lower BDNF levels are associated with an increased risk of incident stroke and with poor prognosis after stroke while elevated levels were associated with decreased risks of major disability and death.(11,12) Aerobic exercise has been shown to increase serum BDNF in the healthy population.(6,13) Whilst being a recommended therapy also during the early subacute phase in many clinical stroke guidelines worldwide aiming to to facilitate neuroplasticity,(14,15) literature on the effectiveness of aerobic exercise on BDNF levels in stroke patients is conflicting. Most evidence is derived from preclinical experiments and only mostclinical studies measuring serum BDNF have a high risk of bias.(16–18)

In the randomized-controlled ‘Physical Fitness Training in Patients with Subacute Stroke’ (Phys-Stroke) trial,(19) BDNF was measured at four study visits up to six months post-stroke. BDNF was alsomeasured after a four-week aerobic exercise intervention in half of the study population (intervention group). In this sub-analysis, we aimed to investigate overall serum BDNF levels over time and to examine potential effects of an aerobic fitness training intervention on BDNF levels.

## Materials and Methods

Here we present secondary analyses of data from the Phys-Stroke trial. The Phys-Stroke trial was a multicentre, endpoint-blinded, randomised clinical trial in which we tested the efficacy of a four-week treadmill-based aerobic training against relaxation therapy as an active control group in patients 5 – 45 days after stroke. The trial was conducted from 26 September 2013 to 30 April 2017 and registered at clinicalTrials.gov (clinicalTrials.gov identifier NCT01953549). Its primary efficacy endpoints have been reported previously.(19) The trial was approved by the local ethics committee of the Charité University Medicine Berlin (EA1/138/13) and patients gave written informed consent.

### Patients and study design

Patients were eligible if they were within the early subacute phase after stroke confirmed via imaging and were moderately to severely affected operationalized by the Barthel Index less or equal to 65. Major exclusion criteria were bleeding from ruptured aneurysm or arteriovenous malformation, severe cardiac or neurologic comorbidities or major psychological or depressive disorders. In- and exclusion criteria are listed in Table 1. All patients admitted to any of seven rehabilitation wards in the Berlin area, Germany, were assessed and subsequently 200 patients were included. After baseline assessment, participants were randomized between an experimental condition (training) and an active control group (relaxation) using an online randomization tool stratified for age and functional impairment.(20) The intervention phase was targeted to deliver 20 sessions over a period of four weeks with a core intervention time of 25 min.

**Table 1:** Inclusion and exclusion criteria.

| **INCLUSION CRITERIA** |  |
| --- | --- |
| 1. | Diagnosis of stroke (inclusion within 5-45 days after stroke onset); ischemic or haemorrhagic (cortical, subcortical, brainstem), as determined by initial MRI/CT scan of the brain) |
| 2. | Age ≥ 18 years |
| 3. | Able to sit unsupported (i.e. without holding onto supports such as the edge of the bed), with feet supported, for at least 30 seconds |
| 4. | Barthel-Index ≤ 65 at inclusion |
| 5. | Considered able to perform aerobic exercise, as determined by responsible physician |
| 6. | Provision of written informed consent |
| **EXCLUSION CRITERIA** |  |
| 1. | Patient considered unable to comply with study requirements |
| 2. | Stroke due to intracranial haemorrhage primarily due to bleeding from ruptured aneurysm or arteriovenous malformation |
| 3. | Progressive stroke |
| 4. | Unable to perform the required exercises due to a) medical, b) musculo-skeletal, or c) neurological problems (for details see below, 4a-c) |
| 4a. | Medical problems: unstable cardiovascular condition, or other serious cardiac conditions (e. g., New York Heart Association criteria for Class IV heart disease, hospitalization for myocardial infarction or heart surgery within 120 days, severe cardiomyopathy or documented serious and unstable cardiac arrhythmias) |
| 4b. | Musculoskeletal problems: restricted passive range of motion in the major lower limb joints (i.e. an extension deficit of $> 20^\circ$ for the affected hip or knee joints, or a dorsiflexion deficit of $> 20^\circ$ for the affected ankle) |
| 4c. | Neurological problems: severity of stroke-related deficits |
| 5. | Required help of at least 1 person to walk before stroke due to neurological (e. g., advanced Parkinson's disease, Amyotrophic Lateral Sclerosis, Multiple Sclerosis) or non-neurological co-morbidities (e. g. heart failure, orthopaedic problems) |
| 6. | Life expectancy of less than 1 year as determined by responsible physician |
| 7. | Drug or alcohol addiction within the last six months |
| 8. | Significant current psychiatric illness defined as medication-refractory of bipolar affective disorder, psychosis, schizophrenia or suicidality. |
| 9. | Current participation in another interventional trial |

MRI = magnetic resonance imaging | CT = computer tomography

Patients in the training group (n = 105) received treadmill-based aerobic exercise over a period of on average 16 (SD 6) sessions with an average of 21 (SD 4) min per session with a targeted heart rate of 55 – 65% of their maximum heart rate. In the relaxation group (n = 95), patients were given muscle relaxation therapy after Jacobsen focused on the upper limb and body in on average 17 (SD 5) sessions with a core intervention time of 24 (SD 3) minutes. Detailed descriptions of the interventions have been reported previously and interventions were delivered by different personnel than the outcome assessment.(19)

### Outcomes

Study outcomes were assessed by blinded outcome assessors at baseline, post-intervention (V1), at three (V2) and at six months (V3) post stroke. Baseline characteristics included medical history, stroke aetiology and symptoms. Further, the Barthel-Index as a measure of ability to perform activities of daily living and maximal walking speed on a 10m hallway were conducted at each visit. Here, patients were asked to walk as fast but safe as possible. Participants are usually considered non-impaired if they score higher than 26 on a 30 points scale. We asked patients for depressive symptoms using the Center for Epidemiological Studies – Depression (CES-D) scale which is a validated tool to be used in the stroke population.(21) Depression has been shown to negatively impact serum BDNF levels before.(10) The CES-D is a 20-item tool in which scores range from 0 to 60, with higher scores indicating greater depressive symptoms. A lie criteria rates validity of the provided answers. We used ≤ −28 as a cut off and imputed data for the analysis with a value below the cut off. Fasting blood samples were collected from all participants at each respective visit, centrifuged and consequently stored at −80° C until analyses of BDNF using Bioplex-Luminex assay (Millipex® MAP Human Neurodegenerative Disease Magnetic Bead Panel 3, HNDG3MAG-36K; Millipore). Bioplex-Luminex assay was used according to the manufacturer’s guidelines. In addition, we assessed several additional biomarkers that have been reported before. For this analysis, we used platelet counts as they strongly correlate with serum BDNF.(22)

### Statistical analyses

Descriptive analyses are presented with mean (SD) or median [IQR] where appropriate. Pearson correlations between levels of serum BDNF and platelet counts were calculated to see whether platelet counts were connected with levels of serum BDNF. Under the assumption of data missing at random, missing values were imputed using imputation with chained equations with iterations of 1000 imputations (MICE package). Analyses were pre-defined but exploratory in nature and therefore no adjustments for multiple comparisons were added (Statistical analysis plan of the Phys-Stroke trial can be found at https://doi.org/10.6084/m9.figshare.5375026.v1).(23) We performed sensitivity analyses on data from all patients without imputed data.

At first, we assessed BDNF progression over time to elucidate its overall progression based on the time of inclusion after stroke. Here, we fitted repeated measures linear mixed models with changes in serum BDNF as the primary outcome and days from stroke to start of intervention clustered in 5 to 15 days, 16 to 30 days and 31 and more days as the explanatory variable. Models were controlled for baseline values and adjusted for age, sex, stroke severity operationalized with the National Institute of Health Stroke Scale (NIHSS), platelet counts, depressive symptoms at baseline and smoking status prior to stroke.(24) Centre was added to account for participants being clustered in different study sites. Different from the primary efficacy analyses published previously, we did not add Centre as a random intercept as this would have resulted in convergence problems due to the high number of covariates but formed three clusters with the early-rehabilitation wards (2 sites) and the geriatric ward being one cluster each and all inpatient rehabilitation wards clustered together (4 sites).

We further wanted to investigate whether allocation to either of the two treatment arms influenced serum BDNF progression. Another repeated measures linear mixed model was modeled where treatment was used as an explanatory variable. Further, a separate model was built with an interaction term of sex and treatment. This was done to explore whether a sex and treatment interaction found during the primary efficacy analyses was also present in serum BDNF. Here, we previously reported an exploratory finding that females interacted with treatment allocation in the sense that females did improve in maximal walking speed in the training group compared to males.

Previous reviews suggested a dose-response relationship of aerobic exercise on BDNF levels. Therefore, we fitted another model with number of intervention days as an exploratory variable. Covariates remained the same throughout all linear models.

Lastly, we wanted to investigate whether serum BDNF levels would be connected with possible recurrent events. Here, we looked whether BDNF at baseline was associated with only the serious adverse events recurrent cerebro- or cardiovascular event within the study period of six months in line with the safety analyses reported previously.(25)

Analyses were conducted with R statistical software (Version 4.3.1) with the packages tidyverse (Version 2.0.0), and mice (3.16.0).(26) Analyses codes are made freely accessible with the publication.

## Results

Data with consecutive blood draws were available from 113 patients. Reasons for patients not receiving blood draws include dropouts and losses to follow-up at later time points. Missing blood draws occurred if patients were unavailable or discharged early. Data flow is depicted in the flow-chart (Figure 1).

**Figure 1:**
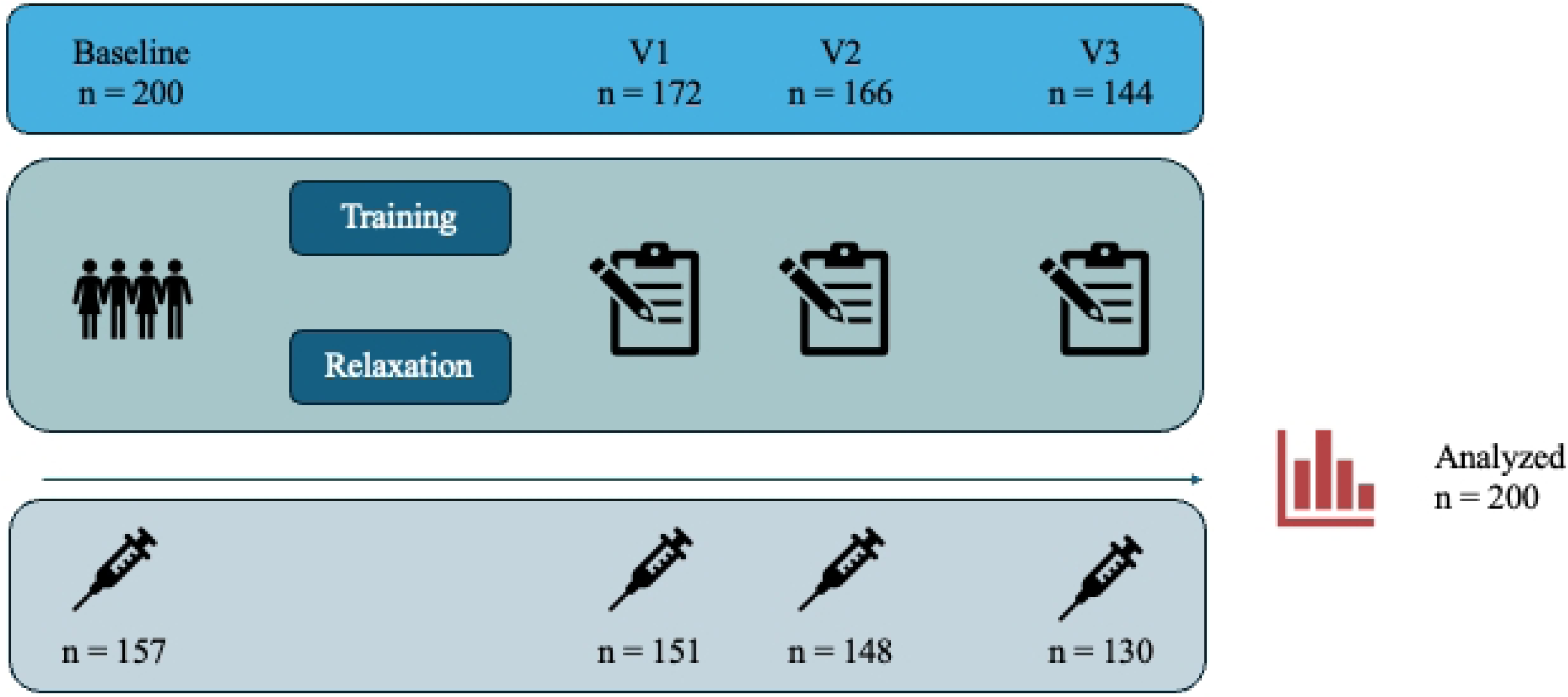
Flow chart of patients with availability of respective blood draws

Descriptive analyses are reported without imputed data. Baseline characteristics from included patients are listed in Table 2. Overall, patients were moderately to severely affected with a median NIHSS score of 8 [5 to 12] and a Barthel-Index score of 50 [35 to 60]. Participants exhibited depressive symptoms at baseline with a median CES-D sum score of 10 [6 to 14]. CES-D data of 24 patients had to be removed and subsequently imputed as they were below the cut-off from the lie criterion. We found a weak positive correlation between serum BDNF at baseline with NIHSS assessed within the stroke unit (*r* = 0.17, *p* = 0.031) and a negative correlation with CES-D values at baseline (*r* = 0.21, *p* = 0.021). We found that participants that presented with concomitant diabetes mellitus (14.2 ng/ml, SD = 9.7, t(146) = 2.4, *p* = 0.019) and cardiovascular disease (13.8 ng/ml, SD = 6.8, t(47) = 2, *p* = 0.052) had overall lower serum BDNF at baseline while those with hypercholesterolaemia presented with increased serum BDNF (19.9 ng/ml, SD = 14.3, t(120) = −2.1, *p* = 0.035) compared to those participants without respective comorbidity. We found no such differences for any of the other comorbidities arterial hypertension, atrial fibrillation, or previous stroke. Platelet counts and serum BDNF levels at baseline were weakly correlated (*r* = 0.14, *p* = 0.087).

**Table 2:** Baseline characteristics.

| Baseline characteristics | Training (n = 105) | Relaxation (n = 95) | All (n = 200) |
| --- | --- | --- | --- |
| Age, mean (SD) | 69 [12] | 70 [11] | 69 [12] |
| Female, n (%) | 45 (43) | 36 (38) | 81 (41) |
| Days post stroke, mean (sd) | 26 (12) | 27 (17) | 27 (15) |
| Ischemic stroke, n (%) | 91 (87) | 90 (95) | 181 (91) |
| Treatment with alteplase, n (%)<sup>#</sup> | 34 (37) | 27 (30) | 61 (34) |
| Previous stroke, n (%) | 27 (27) | 27 (28) | 54 (27) |
| Diabetes mellitus, n (%) | 32 (31) | 31 (33) | 63 (32) |
| Cardiovascular disease, n (%) | 11 (11) | 18 (19) | 29 (15) |
| Hypercholesterolaemia, n (%) | 43 (41) | 37 (39) | 80 (40) |
| Current smoker, n (%) | 31 (30) | 32 (34) | 63 (32) |
| NIHSS, median [IQR] | 9 [5 to 12] | 7 [5 to 11] | 8 [5 to 12] |
| Maximal walking speed, m/s, mean (sd) | 0.4 (0.4) | 0.5 (0.4) | 0.4 (0.4) |
| Barthel Index, median [IQR] | 50 [35 to 60] | 55 [35 to 65] | 50 [35 to 60] |
| CES-D, median [IQR] | 10 [6 to 16] (n = 85) | 11 [6 to 14] (n = 72) | 10 [6 to 14] (n = 159) |
| BDNF, ng/ml, mean (sd) | 19.7 (16) (n = 83) | 14.3 (8.2) (n = 74) | 17.2 (13.2) (n = 157) |
| Thrombocytes, no/ml | 279.5 (89.8) (n = 102) | 282.3 (106.9) (n = 94) | 280.8 (98.1) (n = 196) |

The intervention started in the training group two days earlier than in the relaxation group (27 SD = 12 vs. 29 SD = 13). Compared to patients in the relaxation group, patients in the training group had more severe strokes (median NIHSS score 9, IQR 5 to 12 vs 7, IQR 5 to 11) and higher serum BDNF (mean 19.7 ng/ml, SD 16 vs. mean 14.3 ng/ml, SD 8.2, t(125) = −2.7, *p* = 0.007) values at baseline. Overall, treatment fidelity was fair and had been reported previously.(19)

### BDNF over time

Blood draws were conducted on average on day 27 (SD 15) for the baseline visit, on day 59 (SD 32) for the post-intervention visit V1, on day 94 (SD 29) for the three months visit V2 and on day 181 (SD 8) for the six months visit V3. Progression of serum BDNF in the cohort is depicted in Figure 2a. Overall, serum BDNF levels increased until 3 months post stroke (22.6 ng/ml, 95% CI 19.2 to 26) and remained stable until six months post stroke (24.3 ng/ml, 95% CI 20.6 to 27.9). We looked whether the time point of baseline visit after stroke made a difference in the slopes of serum-BDNF progression if start was clustered in the three timeframes 5 to 15 days (50 participants), 16 to 30 days (68 participants) and 31 and more days (75 participants) but were overall not able to detect large differences (Figure 2b). In general, serum BDNF progressed from 17.8 ng/ml [95% CI 12.7 to 22.8] in the earliest, 18.5 ng/ml [95% CI 15.2 to 21.8] in the middle and 14.6 ng/ml [95% CI 10.9 to 18.4] in the latest group to 23.7 ng/ml [95% CI 16.4 to 30.9], 25.5 ng/ml [95% CI 20.6 to 30.3] and 22.9 ng/ml [95% CI 17.5 to 28.3] respectively at six months post stroke.

**Figure 2:**
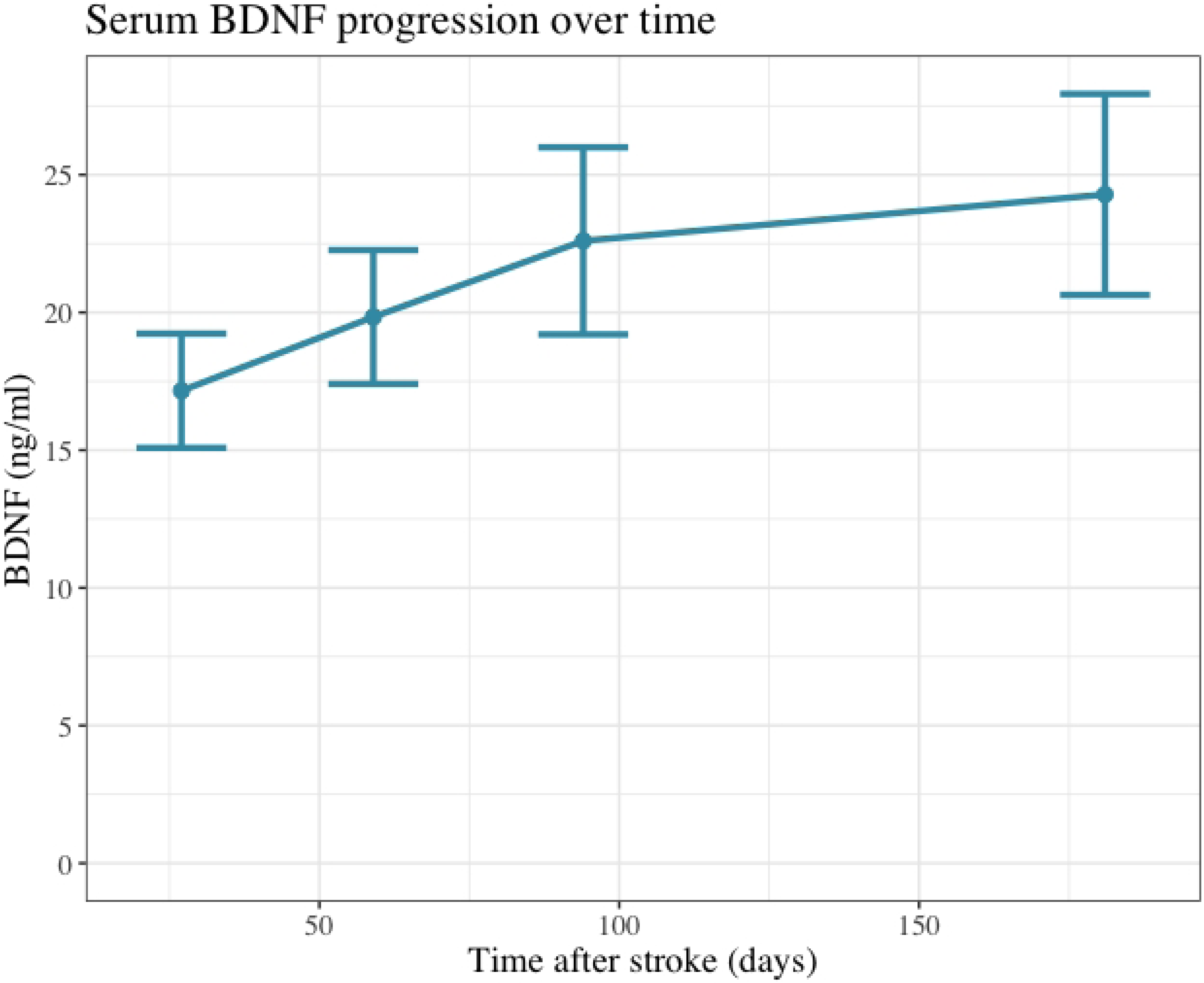
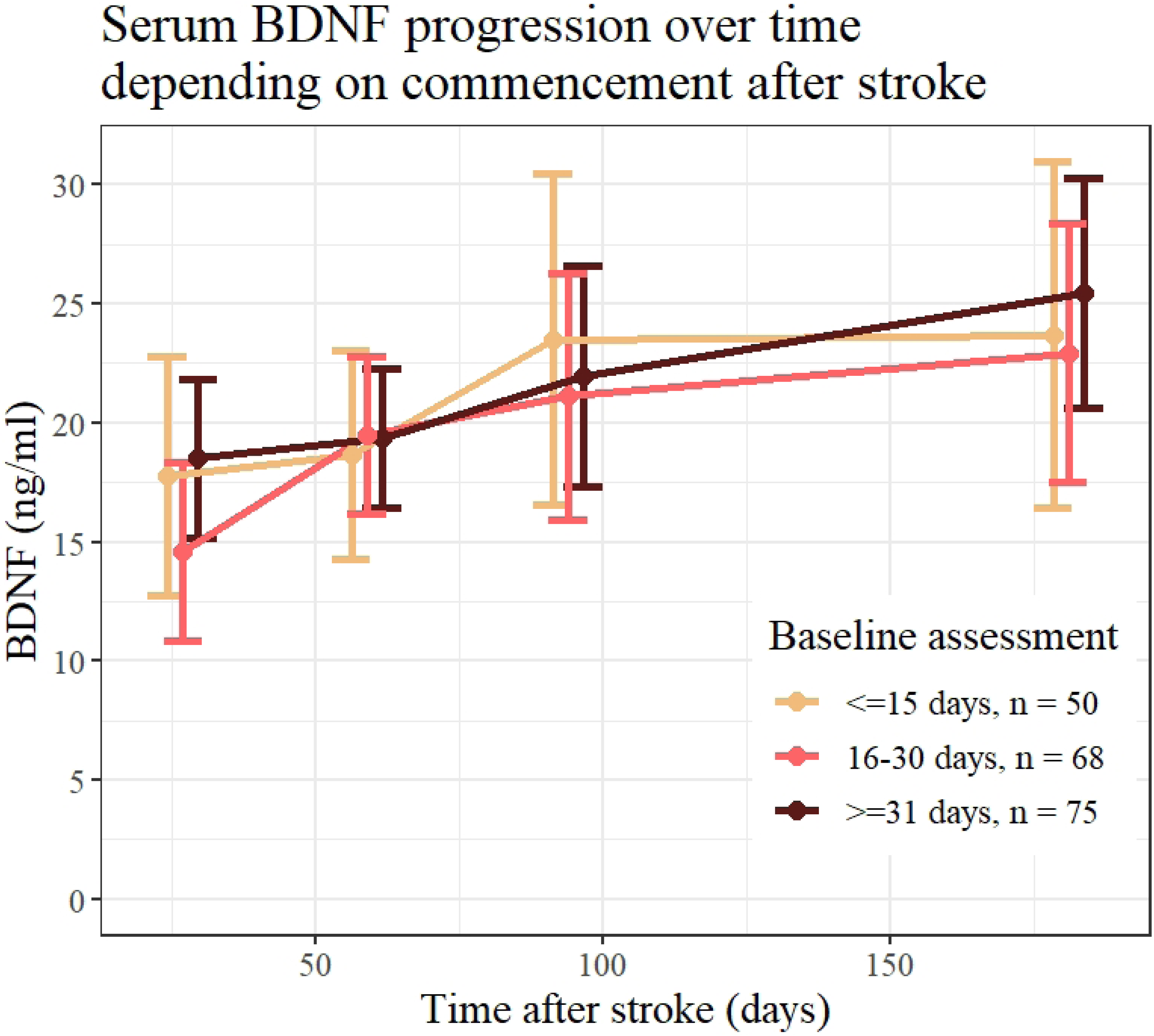
(a) Overall progression of serum BDNF over the study period up to six months post stroke. Time points of assessment are Baseline, after intervention, three months post stroke and six months post stroke. (b) Serum BDNF progression per time window, in which the baseline visit was conducted, clustered into the time frames less and equal to 15 days after stroke, between 16 and 30 days and more than 30 days after stroke.

### Treatment effect

Although patients in the training group started with higher serum BDNF levels, progression was similar between groups and patients in the relaxation group reduced the difference almost completely at six months post stroke (training: 24.6 ng/ml, 95% CI 20.2 to 29 vs relaxation: 24.3 ng/ml, 95% CI 19.8 to 28.8, *p* = 0.95). Also, the changes between individual visits were similar between both groups (Figure 3a).

**Fig 3:**
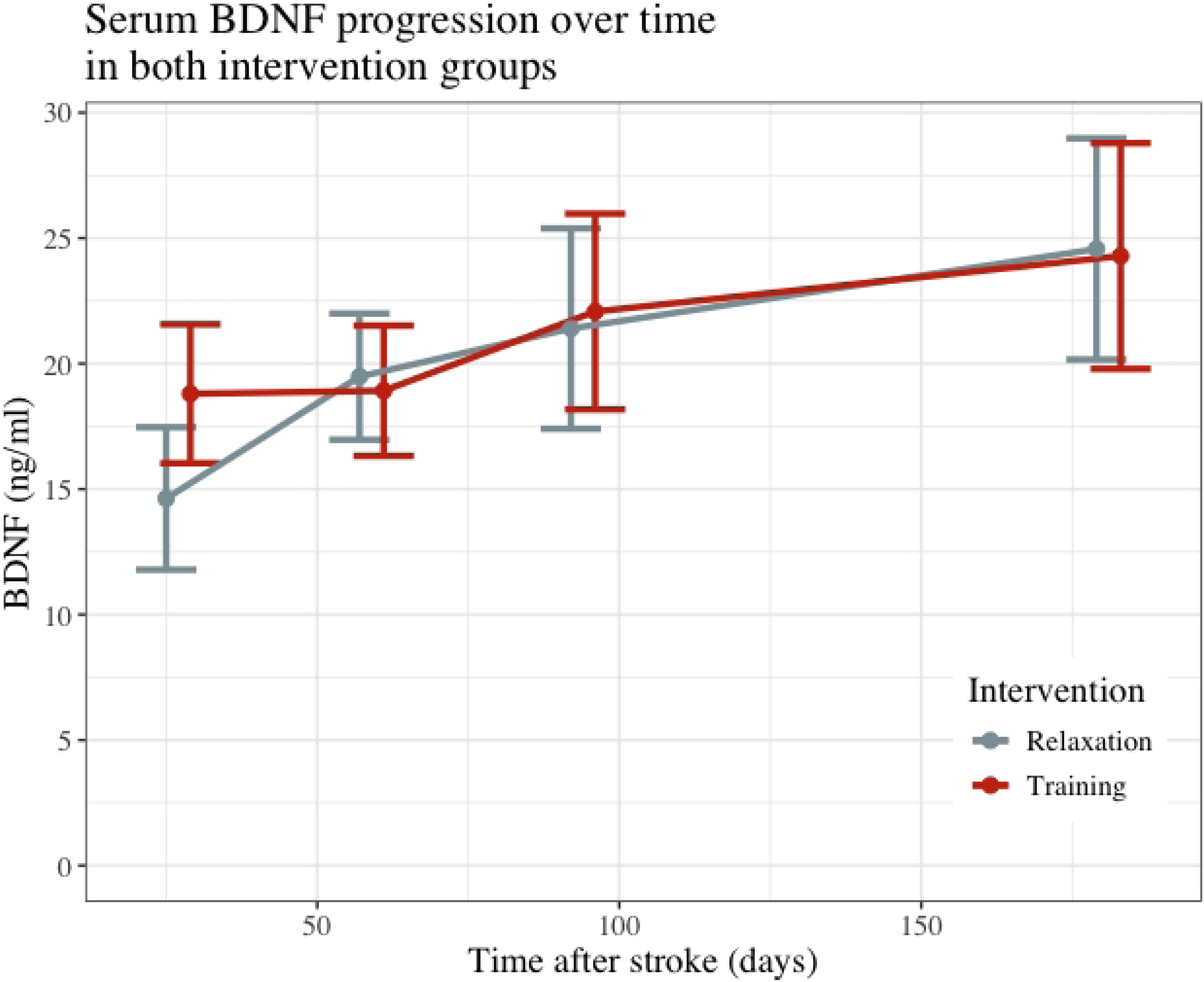
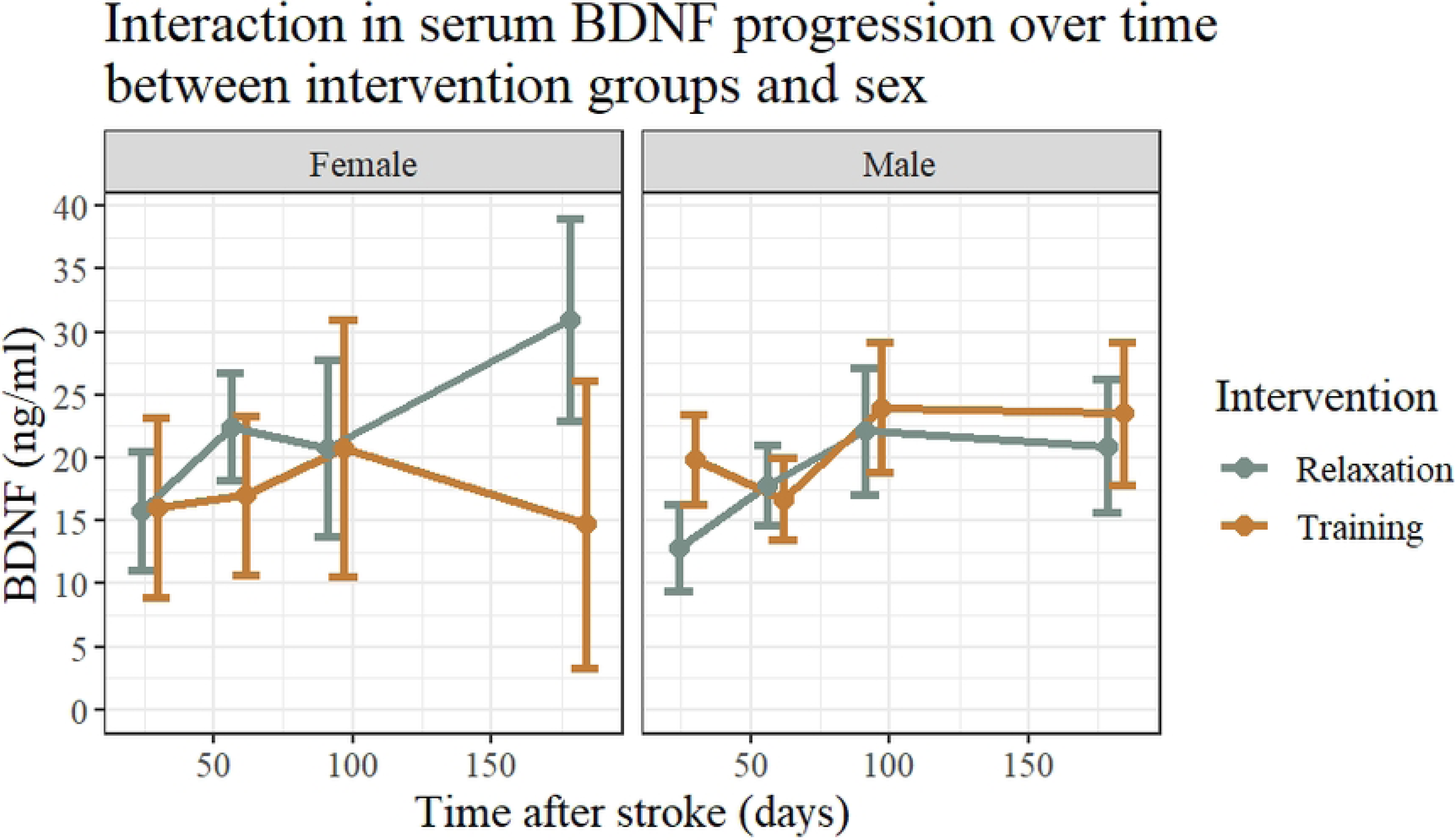
(a) Serum BDNF progression from baseline up to six months post stroke per intervention arm. Time points of assessment are Baseline, after intervention, three months post stroke and six months post stroke. (b) Serum BDNF progression per gender in both treatment arms up to six months post stroke. Values are displayed as means with 95% confidence intervals.

In the main analyses, we reported an interaction of treatment with sex in which females gained higher increases in maximal walking speed at 3 months post stroke if allocated to the training group compared to males. Therefore, we analyzed if similar interaction effects could be seen in serum BDNF (Figure 3b). An interaction could be found but dissimilar to what was expected a sex effect was found in the relaxation group in which females displayed higher increases in serum BDNF up to six months post stroke than males (20.9 ng/ml, 95% CI 15.7 to 26.2 (males) vs 31.0 ng/ml, 95% CI 22.9 to 39.0 (females), *p* = 0.039) while no such effects were seen in the training group (23.5 ng/ml, 95% CI 17.9 to 29.2 (males) vs. 14.7 ng/ml, 95% CI 3.4 to 26.1 (females), *p* = 0.148) (Table 3).

**Table 3:** Progression of BDNF in both groups and per sex.

|  | **Training** |  |  | **Relaxation** |  |  |
| --- | --- | --- | --- | --- | --- | --- |
|  | Male, ng/ml<br>(95% CI) | Female, ng/ml<br>(95%CI) | p-value | Male, ng/ml<br>(95% CI) | Female, ng/ml<br>(95%CI) | p-value |
| **Baseline** | 19.8<br>(16.2 to 23.5) | 16.0<br>(8.9 to 23.1) | 0.334 | 12.9<br>(9.4 to 16.3) | 15.8<br>(11.1 to 20.5) | 0.331 |
| **Post-intervention (V1)** | 16.7<br>(13.5 to 20.0) | 17.0<br>(10.6 to 23.4) | 0.935 | 17.8<br>(14.7 to 21.0) | 22.5<br>(18.3 to 26.7) | 0.078 |
| **3-months post stroke (V2)** | 24.0<br>(18.8 to 29.2) | 20.8<br>(10.6 to 31.0) | 0.568 | 22.1<br>(17.1 to 21.1) | 20.7<br>(13.7 to 27.7) | 0.748 |
| **6-months post stroke (V3)** | 23.5<br>(17.9 to 29.2) | 14.7<br>(3.4 to 26.1) | 0.148 | 20.9<br>(15.7 to 26.2) | 31.0<br>(22.9 to 39.0) | **0.039** |
| **V1 – Baseline** | -0.1<br>(-3.3 to 3.0) | 3.9<br>(-6.3 to 14.2) | 0.935 | 1.0<br>(-2.2 to 4.1) | 5.6<br>(1.4 to 9.8) | 0.078 |
| **V2 – V1** | 7.3<br>(2.1 to 12.5) | 3.8<br>(-6.5 to 14.1) | 0.535 | 4.2<br>(-0.7 to 9.3) | -1.8<br>(-8.7 to 5.2) | 0.167 |
| **V3 – V2** | -0.5 | -6.1 | 0.382 | -1.2 | 10.2 | **0.024** |
|  | (-6.6 to 5.6) | (-18.0 to 5.9) |  | (-6.8 to 4.4) | (1.9 to 18.6) |  |

- 1 Interaction effect of treatment allocation and sex on Serum BDNF levels at each study visit and the change between each visit. Values - 2 based on multiple imputation with chained equations. - 3

### Dose-response relationship

Given that we could not find a treatment effect between patients being randomized to the training compared to the relaxation group, we aimed to elucidate whether the number of sessions received had an influence on serum BDNF progression irrespective of the treatment allocation. In an additional model, we used number of intervention sessions as an exploratory variable but could not find any association neither during the intervention period (β 1.7 ng/ml, 95% CI −4.3 to 7.8, *p* = 0.572) nor between either the post intervention visit and three months post stroke (β 0.5 ng/ml, 95% CI -9.3 to 10.2, *p* = 0.925) or between three to six months post stroke (β 2.3 ng/ml, 95% CI −8.6 to 13.2, *p* = 0.681).

Lastly, we investigated whether serum BDNF at baseline would be associated with cardiac- or cerebrovascular events after stroke but could not find an association (OR 1.01, 95% CI 0.97 to 1.04, *p* = 0.741).

## Discussion

In this article, one of our aims was to describe serum BDNF levels after stroke and to report its progression up to six months follow-up in a cohort of subacute stroke patients of one of the largest aerobic exercise randomized controlled trials in this population. Overall, serum BDNF levels after stroke are reduced compared to the healthy population.(10,27) In a cohort of healthy volunteers aged 18 to 70 years, mean serum BDNF levels were reported to be 32.7 ng/ml (SD 8.33) with aged participants showing higher levels (0.33% increase per year of age).(22) Compared to these reference values our cohort presented with lower serum BDNF values approximately one month after stroke as has been reported also in other stroke populations.(12) In concordance with the literature, serum BDNF levels correlated with stroke severity and inversely correlated with depressive symptoms, although correlations were less pronounced here.(10,27) At baseline, we could identify reduced serum BDNF levels in patients with concomitant diabetes mellitus and cardiovascular disease which has been found in individuals with diabetes and linked to cardiovascular disease earlier.(28–32) In mice, it was shown that a high-fat and refined sugar based diet reduces BDNF levels in the hippocampus and impairs learning mechanisms.(33)

In our cohort, progression of serum BDNF levels up to six months post stroke was modest. In a recent review, serum BDNF levels were found to be stable up to one month post stroke and also in healthy participants, serum BDNF levels did not alter within a 12 months period but presented with high variations of BDNF levels between participants.(10,22) Further, we could not identify differences between participants depending on the date of inclusion to the trial.

Due to its stability, BDNF has good potential to show signs of a response to an intervention such as aerobic exercise. The aerobic exercise intervention in the Phys-Stroke trial was partly chosen due to its neuroplastic effect after stroke. We therefore hypothesized to identify some positive association of serum BDNF levels with our training intervention. Dinoff et al. summarized findings of exercise training on resting BDNF concentrations in which an increase of serum BDNF after aerobic exercise was found, although high between-study heterogeneity existed.(13) Limaye and colleagues synthesized findings from aerobic exercise studies in the stroke population.(17) Here, studies showing an effect of aerobic exercise on BDNF levels were considered high risk of bias while studies with low risk of bias showed no effect.

On the contrary, another systematic review found an overall significant effect of aerobic exercise on BDNF in stroke but evidence was derived mostly from animal studies.(18) In Phys-Stroke, we did not identify a treatment effect on serum BDNF. A study by Boyne and colleagues but also others suggest that exercise effects on BDNF is intensity dependent and might occur only after high-intensity exercise.(16,34,35) In fact, increases in BDNF after exercise has been linked to lactate accumulation lately.(36,37) Also, an effect was seen in interventions of longer durations between 20 to 40 minutes.(17) In Phys-Stroke, participants trained on average 21 minutes in an intensity below the threshold of lactate accumulation which might explain why no treatment effect was seen in our cohort. Lastly, it might be that an increase in serum BDNF due to exercise is not detectable later than 24 hours after cessation of exercise as is the case after acute bouts of exercise.(10) In Phys-Stroke, post intervention visits were conducted between 1 to 2 days post intervention. Therefore, it could be that circulating BDNF levels as a response to exercise had already declined.

In concordance with our subgroup analyses of a sex and treatment interaction on our primary efficacy endpoint maximal walking speed, we aimed to investigate whether we would find a similar interaction on serum BDNF levels. To our surprise, we did find an interaction effect but not within the training but in the relaxation group. Women tend to exhibit higher increases over the course of the intervention phase but also long term between three to six months post stroke within our cohort. Sex differences in aerobic exercise after stroke have been reported before with women showing smaller increases as a response to exercise in the healthy population.(37,38) Literature on interaction effects of sex and relaxation therapy on BDNF levels in the stroke population could not be identified.

To elucidate whether longer durations might reveal dose-response relationships, we investigated whether the number of intervention sessions had an effect on serum BDNF levels irrespective of the treatment allocation but could not find any associations. Distribution of received intervention sessions in the Phys-Stroke trial was skewed as participants were targeted to receive 20 sessions.

Stanne and colleagues reported an association of low BDNF values with incident stroke.(12) In our extended safety analysis, we could not link BDNF with occurrence of cardio- or cerebrovascular events.

### Limitations

Although a robust and independent sequence generation procedure was applied, participants in the training group were more severely affected with higher stroke severity and reduced mobility. Serum BDNF levels were elevated in this group, potentially as a response to the more severe strokes which might have limited the possibility to respond to external stimuli. Analyses reported here are adjusted for baseline values and the progression of BDNF levels in females being allocated to the relaxation group show that an upper limit of BDNF level increase has not been reached. Dropout rates were high in the Phys-Stroke trial and additionally some attrition of BDNF data occurred which could result in a biased effect estimate as most likely severely affected participants were less likely to come to follow-up visits. High dropout rates are common in trials in subacute stroke.(39) We therefore applied robust statistical methods to simulate overall effect estimates including data from all 200 participants. Still, in a study by Naegelin and colleagues, it was suggested in a power analysis that group sizes of 60 participants are already needed to find a change of 20% in serum BDNF levels.(22) It therefore might be that the Phys-Stroke trial was underpowered to assess a positive effect. The Phys-Stroke trial was powered to detect a meaningful change in maximal walking speed and Barthel Index and therefore the results presented here can be seen only as hypothesis generating.

We recruited patients within a time window of five to 45 days post stroke in which recovery processes have already started. Therefore, our participants were most likely in different neuroplastic states when they entered the study and might have showed different progressions of BDNF and intervention responses. We did analyze serum BDNF levels in clusters of time after stroke and could not identify differing progression behavior. Also, participants were almost equally distributed between the three clusters.

Serum BDNF levels have been reported to be influenced by a single nucleotide polymorphism on the BDNF gene. Those Val66Met polymorphism carriers are reported to present with altered activity dependent release of BDNF which may have impacted the effect of exercise on BDNF levels.(13,28,40) In our trial, we did not assess the percentage of Val66Met polymorphism carriers and are therefore not able to estimate its modifying effect on our results.

Lastly, there has been critique on the validity of serum BDNF assessment as a proxy measure to BDNF in the brain. Béjot and colleagues could not correlate serum nor plasma BDNF levels to brain BDNF levels at any time point after stroke in a rat model of stroke.(41) Further, as there is no data on circulating BDNF levels prior to stroke we do not know whether reduced BDNF levels found in stroke patients are a response to the incident or rather an effect on systemic diseases such as diabetes mellitus and cardiovascular diseases as BDNF is normally produced also in other organs and muscle tissue. So far, it is not straight forward to assess brain BDNF values in human subjects. Therefore, circulating BDNF seems currently to be the most appropriate proxy of BDNF levels within the cortex.

### Conclusion

We found overall serum BDNF levels to be low after stroke with moderate increase within the first three months. In our sample, aerobic training did not influence BDNF levels.

## Acknowledgements

## Data Availability

Related data to the primary efficacy endpoints have been published previously. Analysis scripts unique to the current manuscript are freely accessible on github: https://github.com/Funkstille1011/BDNF

## Acknowledgements

We thank all participants for their participation. In addition, we would like to thank Jadranka Denes for oversight of blood sample logistics. The Phys-Stroke study group consists of Ulrike Grittner, Holger Bläsing, Anna Gorsler, Darius G Nabavi, Heinrich J Audebert, Fabian Klostermann, Ursula Müller- Werdan, Elisabeth Steinhagen-Thiessen, Andreas Meisel, Matthias Endres, Stefan Hesse, Martin Ebinger, and Agnes Flöel.

## Author contribution statement

TR has written the first draft and analyzed the data, LD has designed the analysis and analyzed the data, KN analyzed the data and implemented the multiple imputation, AN designed and oversaw the analysis. All authors contributed to the final version of the manuscript and had access to the full data.

